# Discovery of plasma protein biomarkers related to Alzheimer’s disease, sex and APOE genotype

**DOI:** 10.1101/2020.04.15.20066647

**Authors:** Scott B. Laffoon, James D. Doecke, Anne M. Roberts, Jennifer A. Vance, Benjamin D. Reeves, Kelly K. Pertile, Rebecca L. Rumble, Chris J. Fowler, Brett Trounson, David Ames, Ralph Martins, Ashley I. Bush, Colin L. Masters, Paul A. Grieco, Edward A. Dratz, Blaine R. Roberts

**Affiliations:** Florey Institute of Neuroscience and Mental Health and The University of Melbourne Dementia Research Centre, Parkville, Victoria 3010 Australia; Department of Chemistry and Biochemistry, Montana State University, Bozeman, MT 59715 USA; Cooperative Research Centre for Mental Health, Carlton South, VIC, Australia; Australian e-Health Research Centre, CSIRO and Cooperative Research Centre of Mental health, Royal Brisbane and Women’s Hospital, Brisbane, QLD, 4029, Australia; AIT Bioscience, 7840 Innovation Blvd., Indianapolis, IN 46278, USA; School of Medical Sciences, Edith Cowan University, Joondalup, WA, Australia; Department of Biomedical Sciences, Macquarie University, Norht Ryde, NSW, Australia; Department of Biochemistry, Department of Neurology, Emory School of Medicine, Atlanta, GA 30322 USA

**Keywords:** Proteomics, 2D-PAGE, Alzheimer’s disease, biomarkers, plasma, APOE, Zdye, blood

## Abstract

The Australian Imaging and Biomarker Lifestyle (AIBL) study of aging is designed to aid the discovery of biomarkers. The current study aimed to discover differentially expressed plasma proteins that could yield a blood-based screening tool for Alzheimer’s disease. To search for biomarkers and elucidate mechanisms of AD, we immuno-depleted the most abundant plasma proteins and pre-fractionated the remaining proteins by HPLC, prior to two-dimensional gel electrophoresis. The relative levels of approximately 3,400 protein species resolved on the 2D gels were compared using in-gel differential analysis with spectrally resolved fluorescent protein detection dyes (Zdyes™). Here we report on analysis of pooled plasma samples from an initial screen of a sex-matched cohort of 72 probable AD patients and 72 healthy controls from the baseline time point of AIBL. We report significant changes in variants of apolipoprotein E, haptoglobin, α1 anti-trypsin, inter-α trypsin inhibitor, histidine-rich glycoprotein, and a protein of unknown identity. α1 anti-trypsin and α1 anti-chymotrypsin demonstrated plasma concentrations that were dependent on *APOE* ε4 allele dose. Our analysis also identified an association with the level of Vitamin D binding protein fragments and complement factor I with sex. We then conducted a validation study on individual samples using a targeted LC-MS/MS assay. This study indicates that a peripheral protein signature has potential to aid in the characterization of AD. We also found significant associations of protein levels with APOE genotype, indicating that differences in genotype influence the circulating abundances of proteins other than ApoE.

## Introduction

Alzheimer’s disease (AD) is the most common cause of dementia, accounting for 60-80% of cases. It is estimated that there are over 40 million people suffering from AD dementia, with the number expected to rise to over 100 million by the year 2050 (1). More than 75% of those afflicted with dementia remain undiagnosed (2). While a small portion of AD cases are familial, 95% of AD cases are late onset or “sporadic”, being of unknown and possibly heterogeneous etiology (3).

Currently, the diagnosis of AD is made by clinical assessments of cognitive impairment, with the exclusion of diagnosis for other dementias. With the addition of brain imaging techniques, the accuracy of diagnosis is approximately 90% (4). The gold standard for the pre-mortem diagnosis of AD involves positron emission tomography (PET) imaging of amyloid load in the brain (5). Due to the expense, low availability, and the technically challenging nature of PET imaging it is not suitable for population screening and there is a large research effort to develop a peripheral screening tool that can be used to detect those at risk of developing AD. Investigations of cerebrospinal fluid (CSF) biomarkers are proving to be effective for early detection of AD risk(6,7). However, the collection of CSF is not a widespread practice, as it requires an invasive lumbar puncture, as compared with much less-invasive blood tests. This situation has led to an intensive search for blood based biomarkers that may reflect changes in AD that are systemic, or due to either the disruption of the blood-brain barrier (BBB) (8,9) or the trafficking of proteins between the brain and blood(10). While AD is conceptually a brain disease, there is evidence of altered protein expression in the periphery. This has been shown in several avenues of research, including AD related morphological, chemical and proteomic changes in red-blood cells (11-14), alterations of motility in white blood cells and changes in membrane fluidity in leukocytes (15-17), as well as reports of plasma biomarker panels (18-25). Previous proteomics studies have detected significant changes in proteins found in plasma, serum and CSF in AD. However, none of these studies have yielded accurate, specific and reproducible diagnostic markers for AD (26-29). This remains a significant challenge in the field, but there are indications that a blood–biomarker is possible as indicated by recent reports measuring the amyloid beta 40/42 ratio and tau (30-34).

Global discovery investigations of CSF and plasma proteins face an imposing dynamic range of analyte concentrations, spanning approximately 10 orders of magnitude (35,36), whereas the various platforms for proteomic analysis of intact proteins are limited to the most abundant 3-5 orders of magnitude. With the immense heterogeneity of the immunoglobulins, plasma comprises many tens of thousands of protein species, of which approximately 50% by weight is serum albumin (37) and 92% of the plasma protein is composed of 14 of the most abundant proteins (38). Strategies that target the abundant species for removal are designed to bring less abundant proteins into the three to five logs of analytical purview. Reverse phase high-pressure liquid chromatography (RP-HPLC) provides a means for fractionating these species into simpler subsets of proteins. The study presented here combines immuno-depletion of abundant proteins from plasma, followed by RP-HPLC fractionation, and subsequent two-dimensional gel electrophoresis (2DGE). The advantage that 2DGE has as a proteomic platform is its ability to detect changes in a very wide range of post translationally modified protein isoforms. We also employed a pooling strategy to incorporate a larger population in the study to reduce effects of individual variability and to emphasize changes that are related to disease (39,40). Using recently developed Zdyes (41,42) for covalent labeling and differential in-gel comparison of AD and control plasma proteins, we conducted a proteomic screen of protein isoforms in samples obtain from the Australian imaging and biomarker lifestyle study of ageing (AIBL). We have confirmed several previously observed findings (23,24) and report additional plasma protein signatures of AD that vary with APOE ε4 gene dose and sex. Finally, we attempt to validate the discovered targets in individuals on a mass spectrometry based platform.

## Methods and Materials

### Diagnosis of subjects

The Australian Imaging and Biomarker, Lifestyle (AIBL) study is a dual-site, longitudinal cohort study that integrates neuroimaging, biomarker, neuropsychometric and lifestyle(43). The cohort was divided into three groups; cognitively healthy individuals (healthy controls, HC), and participants diagnosed with AD as defined by NINCDS-ADRDA criteria (44) and confirmed by amyloid PET. *APOE* genotypes were determined as previously described (Hixson and Vernier, 1990). Participant recruitment, cohort design and clinical assessments were previously described (Ellis et al., 2009). Due to constraints in sample throughput we pooled samples from each diagnostic category, the demographic and biometric compositions of these pooled group of samples are shown in Table 1. The demographics and biometric composition of the samples used for the validation of the markers discovered in the pooled samples are provided in Table 3. Ethical approval for this study was provided by The University of Melbourne Human Research Ethics Committee.

**Table 1.** Demographic data for each group of pooled plasma (n=12 individuals per pool, three groups per condition). The p-value was calculated with a two-tailed Student’s t-test between All AD and All controls only. ^*^BMI values from 54/72 AD subjects, 60/72 control subjects. Samples are from the baseline timepoint of the ABIL study.

| Parameter | Female<br>AD | Male AD | Female<br>Controls | Male<br>Controls | All AD | All Controls | p-value |
| --- | --- | --- | --- | --- | --- | --- | --- |
| Age in years | 82.2 (1.2) | 77.0 (1.8) | 69.1 (1.5) | 70.5 (4.7) | 79.6 (3.1) | 69.8 (3.2) | 0.0001 |
| % *APOE* ε4 | 37.5 (4.2) | 43.1 (16.8) | 19.4 (2.4) | 6.9 (6.4) | 40.3 (11.4) | 13.2 (8.1) | 0.0001 |
| BMI (kg/mg<sup>2</sup>)* | 25.2 (1.2) | 24.6 (1.7) | 27.2 (1.7) | 25.9 (0.3) | 24.9 (1.4) | 26.5 (1.3) | 0.06 |
| n | 36 | 36 | 36 | 36 | 72 | 72 |  |

### Plasma collection

Plasma was isolated from 15 mL of whole blood collected in two Sarstedt s-monovette ethylenediaminetetraacetic acid (EDTA) K3E (01.1605.008) 7.5 mL tubes with prostaglandin E1 (PGE1) (Sapphire Biosciences, 33.3 ng/mL) pre-added to the tube (stored at 4°C prior to use). After whole blood was collected from overnight fasted participants by venipuncture it was inverted several times and incubated on a laboratory orbital shaker for approximately 15 minutes at room temperature prior to plasma preparation. The whole blood was then combined into 15 mL polypropylene tubes and spun at 200 × g at 20°C for 10 minutes with no brake. Supernatant (platelet rich plasma) was carefully transferred to a fresh 15 mL tube, leaving a 5 mm margin at the interface to ensure the buffy coat was not disturbed. The platelet rich plasma was spun at 800 g at 20°C for 15 minutes with the brake on. The platelet depleted plasma was then aliquoted into 1 mL Nunc cryobank polypropylene tubes (Thermo Scientific, USA) in 0.25 mL aliquots and transferred immediately to a rack on dry ice and transferred to liquid nitrogen vapor tanks (-178°C) until required for the assays.

### Immuno-depletion and fractionation

The plasma from 12 individuals, selected to be pooled together based on sex and disease status (Figure 1), were thawed on ice for 1 hour. The plasma was centrifuged at 16,500 × g for 2 minutes and 50 µl from each individual was mixed together to generate the final pooled samples. Three independent pools of EDTA plasma were prepared from 12 subjects for each of male AD (mAD), female AD (fAD), male healthy control (mHC) and female healthy control (fHC) groups. Pooled plasma samples were immuno-depleted, using a multiple affinity removal system (MARS) 14 column according to manufacturers’ instructions (MARS-14, 4.6 ⨯ 100mm, Agilent), as described in the supplementary material. The flow-through, low abundance proteins were collected in 15 mL conical vials containing 0.82 g urea (SigmaUltra) and 23 µl glacial acetic acid (EMD Chemicals), for a final concentration of 6M urea and 1% acetic acid. After complete mixing, the low abundant protein fractions were frozen and stored at -80° C until reverse-phase (RP) fractionation. The low abundant material was further fractionated into six sub-fractions, using a C18 column (Agilent high-recovery macro-porous 4.6 mm ⨯ 50 mm) operated at 80 °C (see supplemental material for details). Fractions were lyophilized and re-suspended for labeling, with two spectrally resolved fluorescent dyes (Zdye LLC, green emitting BDR-I-227 and blue emitting JAV-I-187). Both fluorescently labelled samples were mixed and first resolved by isoelectric focusing (IEF) on 24 cm pH 3-11 Immobiline™ Drystrips (GE Healthcare) and then electrophoresed on 11% SDS-PAGE gels in the second dimension. Gels were scanned for fluorescence, using a Typhoon™ Trio scanner (GE Healthcare), and false-color images were produced with ImageQuant software (GE Healthcare). Gel image files were imported into Progenesis SameSpots software (Nonlinear Dynamics) for processing, alignment and differential analysis of the relative amounts of protein species in AD and control pools, after normalizing spot volume intensities by the total spot volume intensity in each image Spot volumes were considered significantly changed if there was greater than a 1.3-fold change between groups and a p < 0.05 with a false discovery rate of 0.05 calculated as described in Storey et al. (45), when comparing the six pools (N=12 per pool) of AD subjects with six pools of healthy controls (HC). Additionally, protein spots were compared within male and female pools between AD and HC groups (three pools each for each sex). When comparing differences between the sexes, a cut-off of greater than 1.7-fold change in protein levels was applied. Spearman’s rank correlation and linear regression was used to investigate the relationship between the number of APOE alleles and ApoE protein quantity.

**Figure 1.**
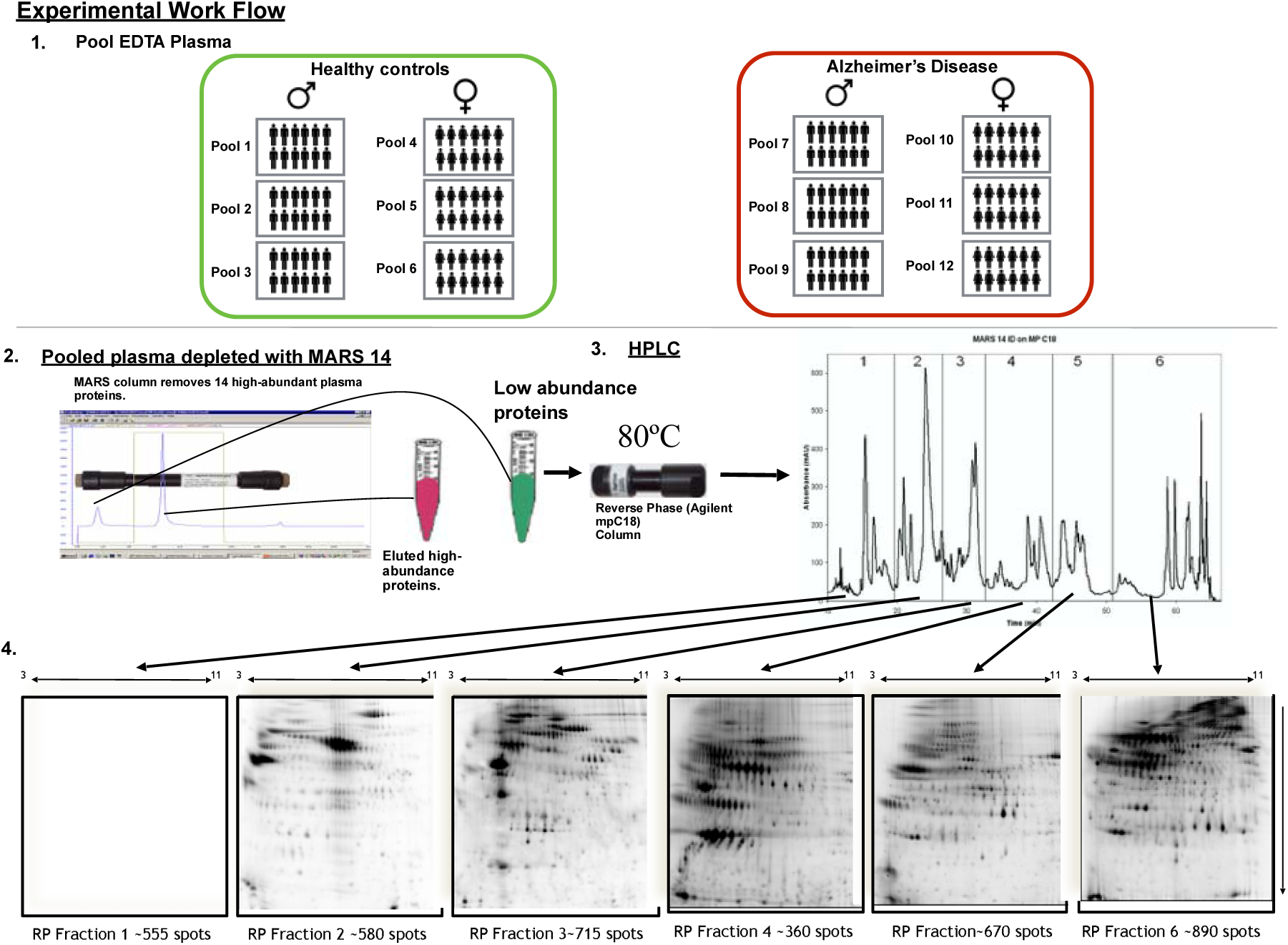
Study design and experimental workflow. (1) Shows how the control and Alzheimer’s disease plasma was pooled into sex specific and disease specific groups. Each pooled group was generated from 12 individuals. (2) The pooled plasma was then depleted of the 14 most abundant proteins using the multiple affinity removal system (MARS14, Agilent Tech.). The low abundance proteins were further fractionated using reverse phase chromatography (3) to generate 6 fractions (4) for 2D differential gel electrophoresis of proteins labeled with Zdyes™(4).

### Identification of proteins-of-interest

To identify changing protein variants, spots-of-interest were excised manually from analytical or preparative gels of fractionated proteins, for in-gel digestion with proteomics grade porcine trypsin (Sigma-Aldrich, Castle Hill, Australia), following previously published methods for in-gel digestion (46). Mass spectrometry data [obtained as described in the supplementary material] were used to search the Swiss-Prot *Homo sapiens* database, using MASCOT version 2.4.1 (Matrix Science). Fixed modification of carbamidomethylation of cysteine, and variable modifications including oxidation of methionine, deamidation of asparagine and glutamine, and a single missed cleavage were allowed for the protein searches, using 2+ and 3+ peptide charge states.

### Selected reaction monitoring QQQ mass spectrometry

Peptides were selected and synthesized, with heavy isotope labeling, for 76 protein targets by MRM-Proteomics (BAK76-kit, Canada). Samples were processed using the manufacturer’s protocol (additional details in supplemental materials). The liquid chromatography mass spectrometry was conducted on with a 1290 UHPLC coupled to a 6495 QQQ mass spectrometer (Agilent Technologies, Mulgrave, VIC).

Statistical analyses were conducted using: Progenesis QI (Waters, UK) for the 2D gel data; GraphPad Prism v 8.0 for Mac OS X (GraphPad Software, USA) for the ROC; t-test analysis: One-way ANOVA with Sidak correction of multiple comparisons and Chi-squared test. A p-value < 0.05 was considered significant.

## Results

Demographic details are listed in Table 1. The AD cohort was significantly older by 9.8 years (AD group 79.6 ± 3.1 years, HC group 69.8 ± 3.2 years, p = 0.0001) and the AD population was significantly enriched in the APOE ε4 allele (40%, AD; 13%, HC; p = 0.0001) as anticipated (47). Body mass index (BMI) data were available for 54/72 AD subjects and 60/72 HC subjects. Using the available data showed a trend (p = 0.06) of somewhat lower BMI in AD pools (24.9 ± 1.4 kg/m^2^) than HC pools (26.5 ± 1.1 kg/m^2^). All of the controls had an amyloid PET scan confirming the lack of significant brain amyloid accumulation and approximately half of 56/72 of the AD cases had a corresponding PET, which confirmed the AD diagnosis. An extensive analysis of the PET data collected on this cohort can be found in Pike et al. (2007) (48).

### Discovery 2D gel investigation of human plasma

The immuno-depletion and RP-HPLC fractionation strategy produced six fractions (F1-6) of proteins for comparison by 2DGE (Figure 1). Representative analytical gel images of each of the six RP fractions are shown in Figure 2. Approximately 3,400 unique protein species were analyzed by this method after reducing the total spot count by 10%, to correct for proteins estimated that eluted in more than one fraction. The inclusion of RP-HPLC fractionation resulted in a roughly linear 6x increase in the quantity of protein spots measured. The increase was attributed to resolution of proteins that co-migrate on 2DGE which have different hydrophobic characters and are thus separated by RP-HPLC, reducing interference in the analysis of gels. In addition, RP-HPLC enriches proteins, allowing lower abundance species to be appear with stronger fluorescent signals on the 2D gels.

**Figure 2.**
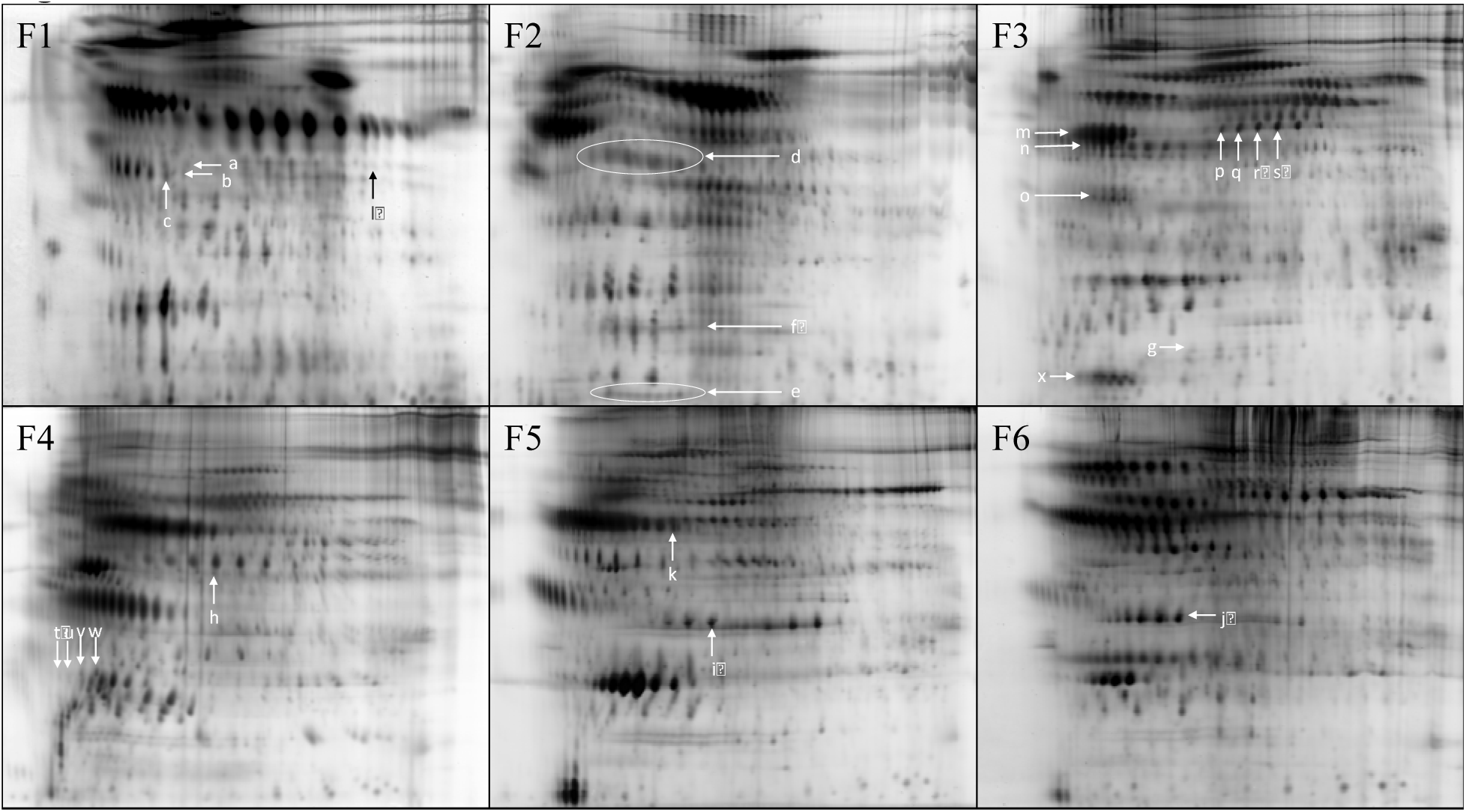
Representative 2D gel images from the six RP fractions. The arrows indicate the protein spots with abundance changes in AD pools compared to HC, as discussed in the Results and Discussion sections.

Spots that met the inclusion criteria for significant differences (>1.3 fold, adjusted p-value <0.05), as described above, are indicated by the arrows in Figure 2 and listed in Table 2. Isoform variants, subunits or cleavage products of eight proteins that discriminated AD from control, according to the inclusion criteria, were identified: zinc α 2-glycoprotein (ZAG), histidine-rich glycoprotein (HRG) fragment, haptoglobin (Hpt), vitamin D binding protein (VDBP), complement factor I (CFI), inter-α trypsin inhibitor (ITHI), α-1 anti-trypsin (α1AT) and apolipoprotein E (ApoE). One significant spot in fraction #1 (F1) was unidentified. Features of proteins that showed significant abundance changes are as follows:

**Table 2.**
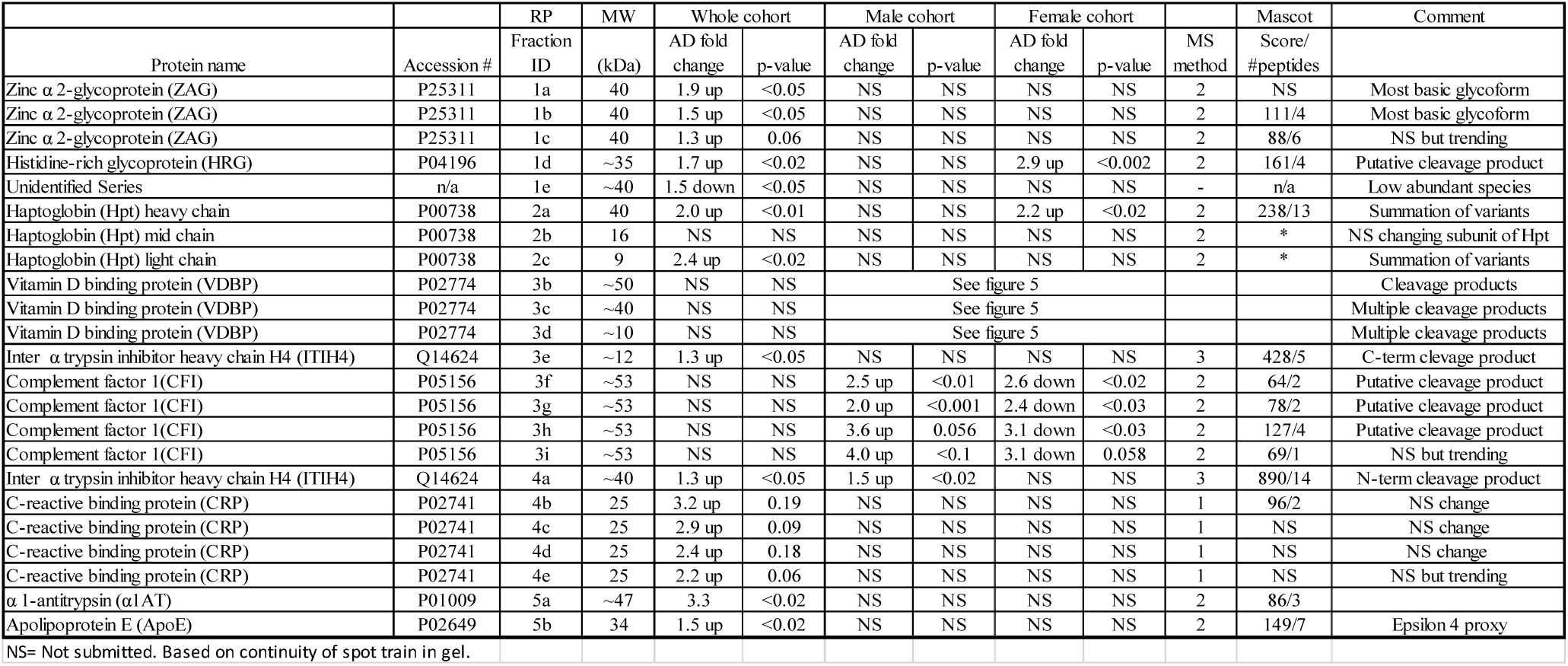
Differentially expressed proteins discovered with 2D gel proteomics with a p-value <0.05.

### Zinc α 2-glycoprotein

Two putative glycoforms of zinc α 2-glycoprotein (ZAG, Figure 2 F1, arrows a and b) showed significant increase in abundance in the AD pools (1.9-fold, p < 0.05 and 1.5-fold, p < 0.05 respectively, Table 2). An adjacent isoform (Figure 2 F1, arrow c) also showed a trend towards being increased in AD pools although it did not quite meet statistical significance (1.3-fold, p = 0.06).

### Haptoglobin

Haptoglobin (Hpt) is an abundant, acute phase response (APR) polymeric glycoprotein that is synthesized in the liver and is composed of varied combinations of four disulfide linked chains (two α^1^ chains of ∼8.9 kDa that differ by a single amino-acid substitution, α^2^ chain ∼16 kDa, and β chain ∼45 kDa). Normal human populations commonly possess quaternary structural variations and heterogeneous stoichiometry of disulfide-linked Hpt chains (49), potentially yielding proteins composed of identical chains with differing pIs. In the current study the disulfide bridges were intact in the first 2D gel separation dimension and then reduced after isoelectric focusing, so the subunits in the second dimension had the same pI patterns as the intact Hpt complexes. To avoid issues arising from mis-matching of heterogeneously focused spots in the comparison between pools, we quantified each of the Hpt spot-trains at 8.9 kDa, 16 kDa and 45 kDa (Figure 2 F2, arrows d, e, and f respectively) as the summed value of the respective constituent isoforms. Two haptoglobin (Hpt) chains (β chain ∼45 kDa - Figure 2 F2, arrow d and α^1^ chain ∼8.9 kDa - Figure 2 F2, arrow e) were significantly higher in AD pools (2.0-fold, p < 0.01 and 2.4-fold, p < 0.02 respectively, Table 2). No significant difference was found in Hpt α^2^ chain (Figure 2 F2, arrow f). False-color image overlays of the F2 gels, qualitatively representing changes in Hpt species, are shown in Figure 3, where the ovals in the upper right image indicate the three different subunit spot trains.

**Figure 3.**
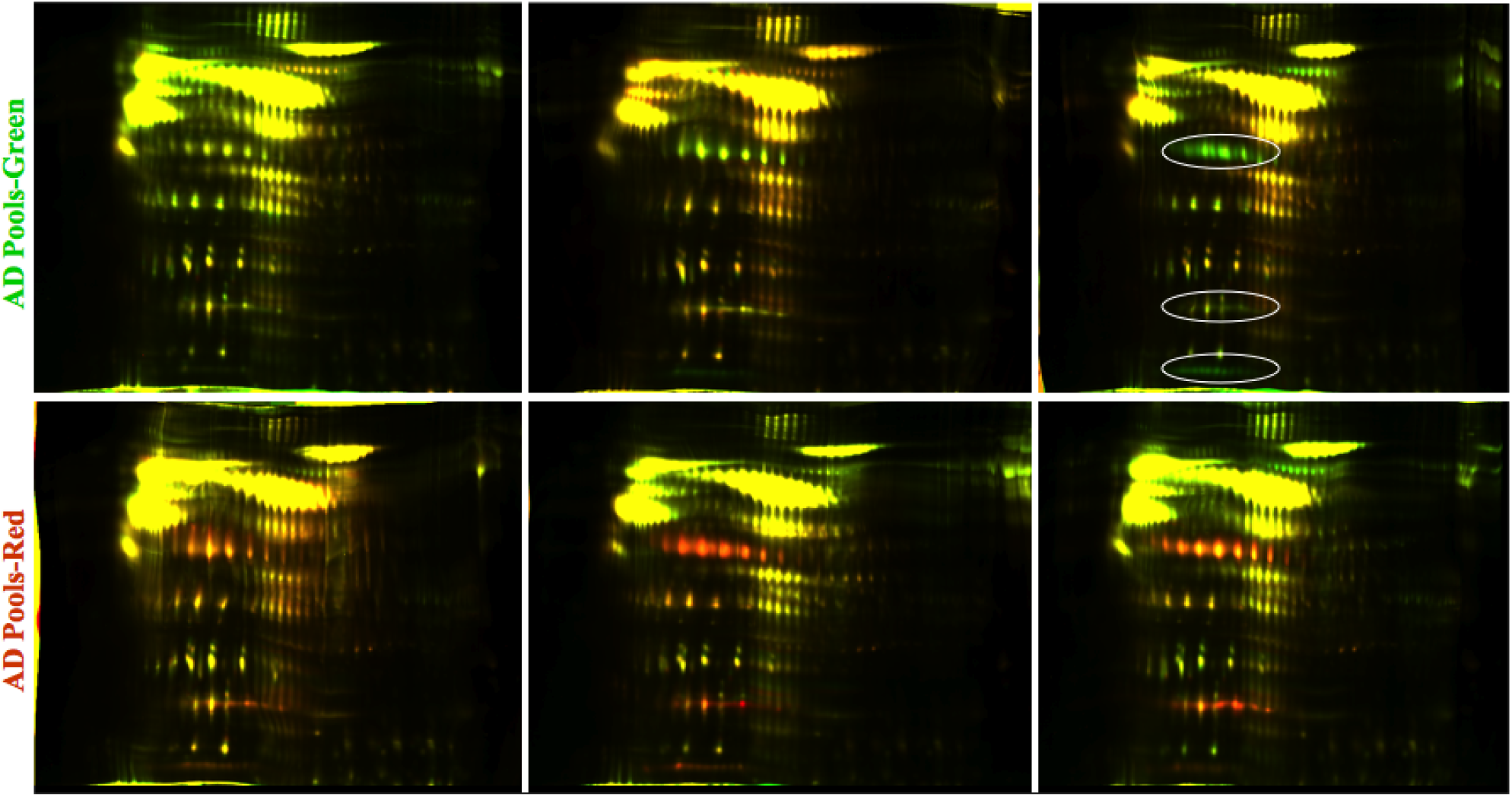
False-color image overlays of F2 two-color multiplex gels prior to spot-matching alignment. Three subunit chains of Hpt are outlined in ovals in the upper right image. The AD pools are represented by green in the upper three images and by red in the lower images, reflecting the dye labeling reversals. Yellow represents approximately equal contribution of intensity from AD and control proteins.

### Inter-α trypsin inhibitor

Two cleavage products of inter-α trypsin inhibitor (ITIH) heavy chain were significantly higher in AD pools. A 12-15 kDa C-terminal cleavage product (Figure 2 F3, arrow g) and an approximately 40 kDa N-terminal cleavage product (Figure 2 F4, arrow h) were both increased 1.3 fold in AD pools (p < 0.05 and p < 0.02 respectively, Table 2).

### Apolipoprotein E

The most chemically basic protein spot of the apolipoprotein E (ApoE) spot train (Figure 2 F5, arrow i) was significantly higher in the AD pools (1.5 fold, p < 0.02, Table 2). The 34 kDa ApoE spot train results from the superposition of the protein products of the APOE ε2, ε3 and ε4 alleles carried by the individuals contributing to the pools. Each of the alleles produces three clearly visible protein spot clusters, likely due to heterogeneous deamidations and glycosylations (50). The substitution of an arginine for a cysteine in ApoE ε4 (C112R), results in a shift of its isoelectric point (pI) toward the cathode, basic (right shift) relative to the ApoE ε3 position. Additionally, plasma proteins are subject to non-enzymatic deamidation *in vivo*, a process that adds negative charges (51), creating features of the horizontal spot trains by shifting species discretely toward the anode, leftward on gels. The partially overlapping superposition of ApoE ε3 with ε4 variants appeared as four clusters of spots in most of the pooled samples, with the most chemically basic cluster comprising only ApoE ε4 variants. Therefore, the most basic cluster of ApoE spots provides a protein isoform proxy for the relative amounts of APOE ε4 allele within the pools. Figure 4A shows strong Pearson correlation (R^2^ = 0.82, p < 0.0001) of the normalized spot intensity of the most basic ApoE “ε4 proxy” variant, with the averaged gene dosage of the APOE ε4 allele in the pools of this study.

**Figure 4.**
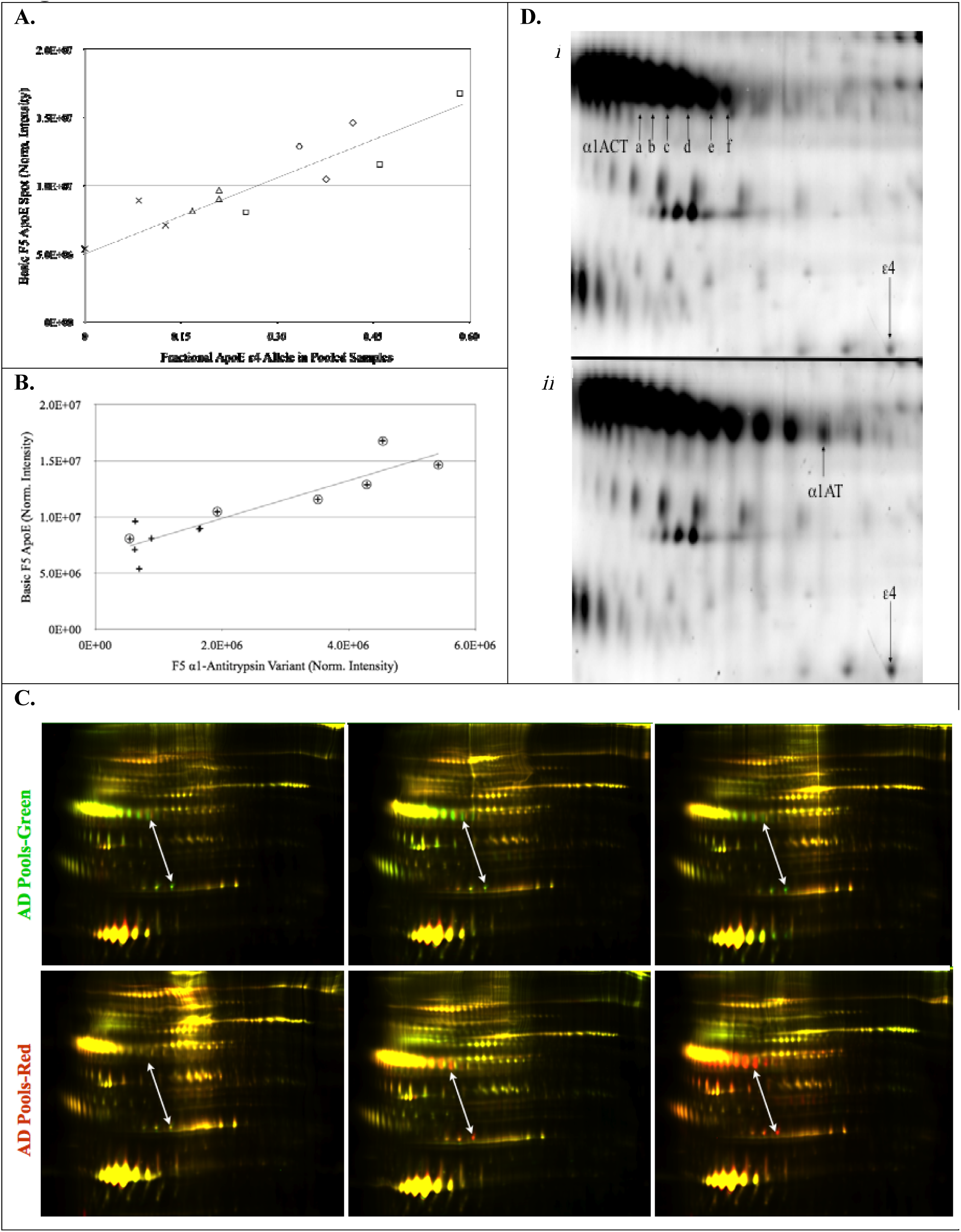
A. Regression analysis of the basic F5 ApoE variant (ε4 proxy) with the fractional *APOE* ε4 gene dosage of the 12 pools: R^2^= 0.83, p< 0.0001. Pool symbols are male healthy controls -╳, female healthy controls -▵., male AD -□, female AD -◇. B. Regression analysis of total cohort, non-binding α1-antitrypsin (α1AT) variant vs. apoE ε4 proxy: R^2^ = 0.82, p< 0.0001; Pools are n=12. Group symbols are: AD - ⍰, HC - +. C. F5 multiplex gels that have not been aligned for spot-matching. Arrows indicate the correlated changes between the ε4 proxy spot (lower end) and the AD significant α1AT variant (upper end). The AD pools are represented by red in the upper three images and by green in the lower images, reflecting the dye reversal. Yellow represents approximately equal contribution of intensity from AD and control proteins. D. Detail from a pair of multiplexed gel images representative of low ApoE ε4 containing pools (panel *i*) and high ApoE ε4 containing pools (panel *ii*). The level of α1ACT isoforms correlated with the 34 kDa ApoE ε4 proxy spot that is shown in lower right-hand corners of the panel *i* and *ii* images. Regression analysis Pearson correlations: a- p=0.012, R^2^=0.45; b- p=0.002, R^2^=0.61; c- p=0.003, R^2^=0.56; d- p=0.002, R^2^=0.61; e- p=0.007, R^2^=0.51; f- p=0.003, R^2^=0.58. None of the α1ACT spots significantly discriminated the AD group from the HC group. The α1AT spot that significantly discriminated AD from HC pools (3.3 fold, p < 0.02,) is shown in panel *ii*.

F6 gels exhibited a series (Figure 2 F6, arrow j) comprising six to seven variants of the same apparent molecular weight (∼35 kDa) as the F5 ApoE series. However, none of the F6 ApoE variants significantly correlated with APOE genotype.

### α-1 Anti-trypsi

A circulating species of α-1 anti-trypsin (α1AT) that was not bound by the immuno-depletion column (Figure 2 F5, arrow k) was significantly higher in AD pools (3.3 fold, p < 0.02, Table 2). The α1AT variant also closely correlated in regression analysis with the ε4 proxy spot volumes (Figure 4B, R^2^ = 0.83, P < 0.0001). The α1AT variant that correlated with the ε4 proxy spot is shown in Figure 4D, where the *i* panel has low ε4 proxy and the *ii* panel has high ε4 proxy. The relationship of these spots is qualitatively represented in the false-color image overlay in Figure 4C, indicated by white arrows.

An unidentified protein spot (Figure 2 F1, arrow l) was significantly lower (1.5-fold, p < 0.05, Table 2) in AD pools.

### Sex specific differences

Vitamin D binding protein (VDBP) cleavage product isoforms, (Figure 1 F3, arrows n, o, × and Figure 5, demonstrated asymmetric sex-dependent AD associations. In female AD pools, several cleavage products of VDBP were in significantly higher concentrations relative to female control pools (p < 0.05). In male AD pools, many of the same VDBP cleavage products had significantly lower concentrations, relative to male control pools (p < 0.05, Figure 5).

**Figure 5.**
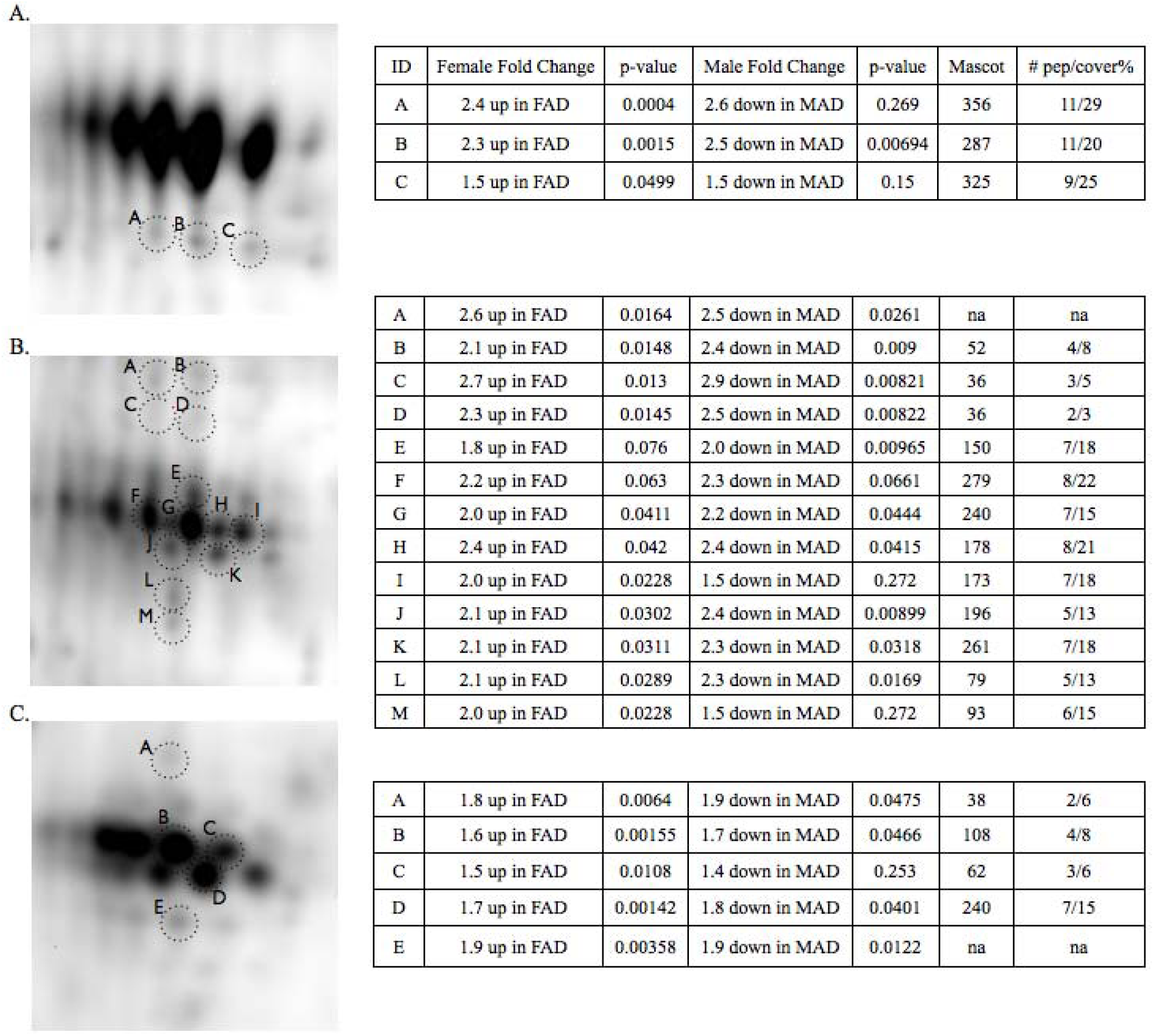
Intact and cleaved VDBP with sex specific changes shown in tables on the right. A. Intact (top spot train) and cleaved VDBP (spots A, B, C). This spot train corresponds to n in Figure 2 F3. The intact VDBP spots were saturated and masked from the Progenesis analysis. B. Cleaved VDBP (A - M). This cluster of cleavages corresponds to o in Figure 2 F3. C. Cleaved VDBP (A - E). This cluster of cleavages corresponds to × in Figure 2 F3. Significant changes (p < 0.05) are shown in bold.

Isoforms of complement factor I (Figure 2 F3 arrows p, q, r, s) also showed significant asymmetrical sex-specific dysregulation in AD (Table 2).

### Targeted Validation of 2D gel biomarkers

The discovery experiment conducted with 2D gels had several limitations, including the use of pooled plasma, which limits the ability to determine the potential for these markers to provide meaningful discriminatory signals in individuals. To validate the differentially expressed proteins in individual samples, we used a multiplexed quantitative LC-MS/MS assay developed by MRM proteomics (BAK76, Canada). Plasma (Li heparin) samples were obtained from AIBL (Demographics Table 3) and included an even number of males and females and cognitively normal controls (n=44) and probable AD (n=44). We could not confirm the elevation of any of the eight proteins we discovered using the pooled samples (Figure 6). However, we did discover that complement C3, beta-2-microglobulin and peroxiredoxin-2, proteins, previously reported to be changed in the literature (29,52-54), were elevated in AD plasma (Figure 6). We did not replicate any of the associations we observed with sex or genotype in the discovery experiment (p >0.05, one-way ANOVA). However, this likely to be due to the inability to distinguish the several proteoforms that were observed by 2D gels with an assay that measures total abundance of all the proteoforms of a protein. Receiver operating characteristic (ROC) analysis for complement C3, beta-2-microglobulin and peroxiredoxin 2 indicate that they only have weak discriminatory (accuracy ranged between 60-65 %) ability to distinguish between cognitively normal from AD (Table 4).

**Table 3.**
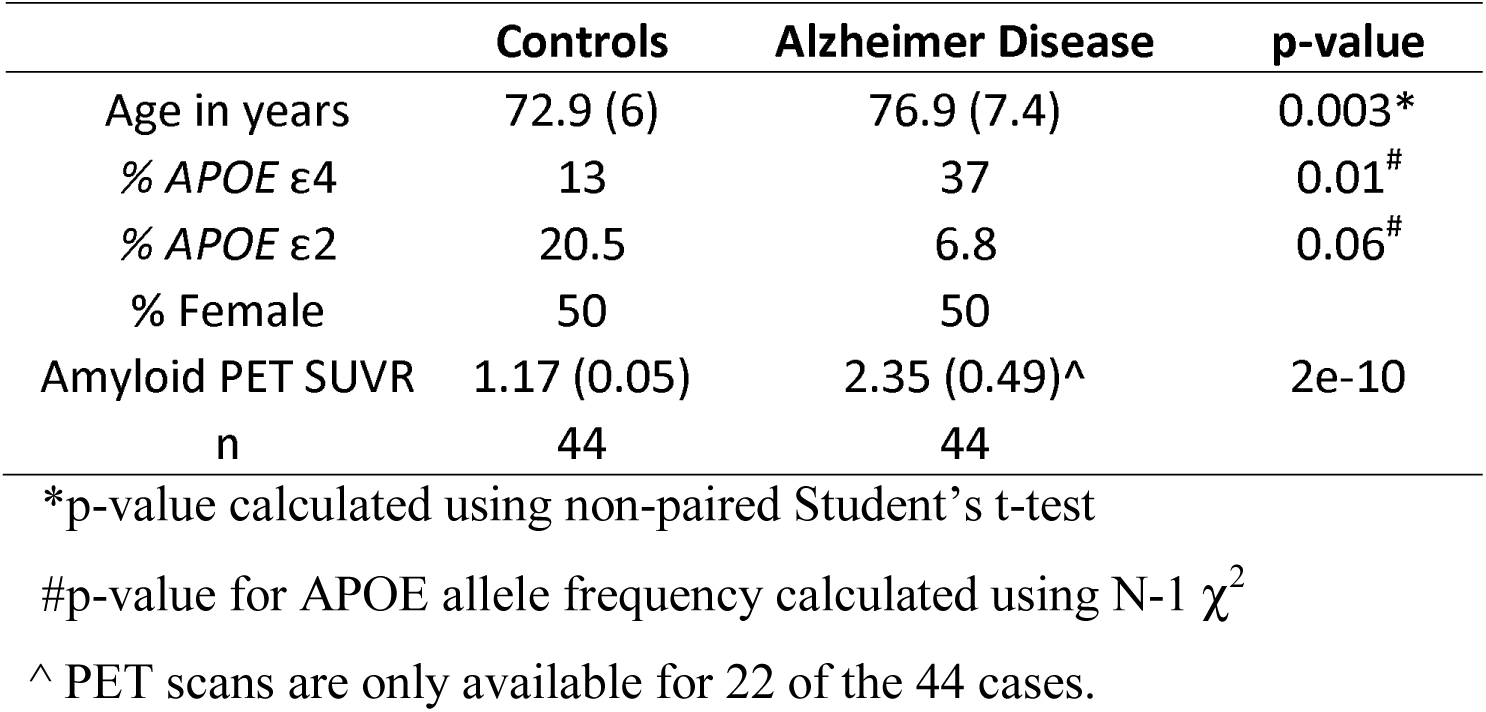
Demographic data for plasma samples used for targeted mass spectrometry biomarker validation.

**Table 4.** Receiver operating characteristic (ROC) analysis of proteins measured by targeted LC-MS assay showing ability to distinguish controls (n=44) from Alzheimer’s case (n=44).

| Protein ID | AUC | Sensitivity % | Specificity % | p-value |
| --- | --- | --- | --- | --- |
| Complement C3 | 0.65 | 69.8 | 59.1 | 0.014 |
| Beta-2-Microglobulin | 0.64 | 47.8 | 80 | 0.022 |
| Peroxiredoxin-2 | 0.60 | - | - | 0.098 |

**Figure 6.**
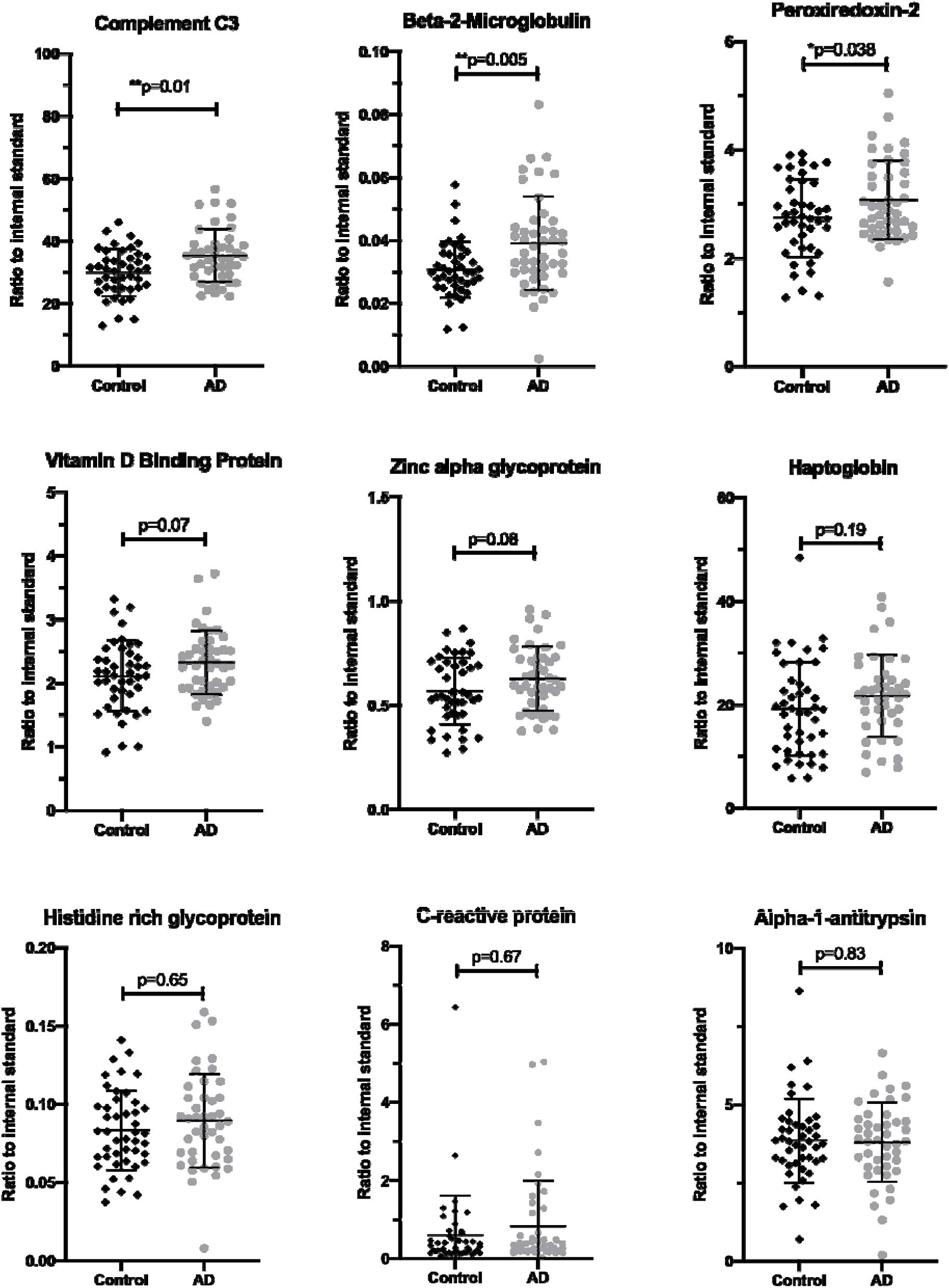
Measurement of plasma biomarkers in control (n=44, black diamonds) and Alzheimer’s disease (n=44, grey circles) plasma. Biomarker candidates discovered using 2D gels were targeted and measured using LC-MS/MS and heavy labeled ([^13^C_6_, ^15^N_2_ -lysine] or [^13^C_6_, ^15^N_4_-arginine) peptides as internal standards. P-values were calculated using a one-way ANOVA and Sidak correction for multiple comparisons.(94)

## Discussion

The effect of the APOE genotype on protein expression is an important consideration for the refinement of discovery and validation of future biomarkers in plasma (55). Consistent with this found significant correlation of increasing α1-antitrypsin (α1AT) with APOE ε4 genotype in plasma. We also found increased levels of cleavage products of VDBP in females with AD and a decrease in the cleavage products in males with AD. (Figure 5). Due to the enrichment of APOE ε4 genotype in AD cohorts, studies that compare controls vs AD are also confounding populations of 20% APOE ε4 positive versus an AD population with 40-80% APOE ε4 positive. To the best of our knowledge the current study is the first plasma proteomics study indicating that α1AT or α1 anti-chymotrypsin (α1ACT) have correlations with the APOE.

α-1 Anti-trypsin (α1AT) is an abundant, circulating, acute phase response (APR) serine protease inhibitor (serpin) that was targeted for removal from the analyte proteins by immuno-depletion (Figure 1 MARS14). The close correlation of α1AT with the ε4 proxy variant, shown in Figure 4B, suggests that α1AT may be a marker of the mode by which APOE ε4 contributes to the etiology of AD. It is of interest that α1AT, which is a serpin with known associations to AD in blood (56-58), CSF(59,60) and senile plaques (61) also correlated with the APOE ε4 allele (Figure 4B), however the specific physiological role of α1AT in tissues is not well elucidated. The Alzgene website shows a single positive genetic association of AD with an α1AT allele, from a small, unconfirmed 1996 study (62). Analysis of total α1AT in neat undeleted plasma from individuals did not display any significant changes (Figure 6).

The APR serpin, α1ACT, did not significantly discriminate AD from control pools, however, regression analysis of α1ACT variants significantly correlated with the ε4 proxy spot volumes (Figure 4D). α1ACT has been reported to have genetic associations with AD (63,64) and has been found in elevated concentration in plasma (65,66) and CSF (67) in AD. α1ACT is more highly expressed in the brains of AD patients and is a component of senile plaques (68), as is ApoE (69). Previous studies have suggested relationships between plasma concentrations of α1ACT and APOE genotype, but the results have been inconsistent. A radial immunodiffusion study found no correlation of APOE genotype with levels of α1ACT in plasma or serum (70). An enzyme linked immuno-sorption assay study, showed α1ACT to have positive predictive value for the rate of cognitive decline in ε4 carriers, and no age-dependence, in AD patients (71). A nephelometric assay study showed higher concentrations of plasma α1ACT in AD patients who were non-APOE ε4 carriers (72). Functionally, α1ACT was shown to induce hippocampus and cerebral cortex plaque formation in mouse models of AD (73), as well as the phosphorylation of tau in the neurons of transgenic mice (74). Both α1ACT and α1AT bind Aβ and suppress its *in vitro* fibrillization, which is a necessary process for the formation of senile plaques *in vivo* (75). Thus its role in AD may be more important at the functional level then as a maker of disease.

The elevated Hpt that we observed along with other circulating APR proteins may be a peripheral response to activated endothelia of the brain vessels. Various means by which cross-talk between central and peripheral inflammation may occur have been summarized (76). The possibility that Hpt flow-through from the immuno-depletion column enriched a sub-population of non-binding conformers may be supported by the selective enrichment of 45 kDa and 8.9 kDa chains in the significantly changed species of AD pools, without a significant change of the 16 kDa chain. The question of whether Hpt was significantly higher in the flow-through because it exceeded the binding capacity of the column, or because of steric hindrance was beyond the scope of this study. In agreement with our observation, a previous report using a MARS column also observed an increase in Hpt in both AD and type 2 diabetes plasma (77) indicating that an Hpt increase is likely due to a general increase in inflammation. When measured in individuals there was no significant increase (Figure 6) in Hpt but this may be due to the inability of the LC-MS/MS method used to discriminate between the 45, 16 and 8.9 kDa chains.

Systemic inflammation is commonly measured clinically by assay of plasma C-reactive protein (CRP) (78). Circulating CRP is primarily synthesized in the liver and, like Hpt, CRP synthesis is mediated by the pro-inflammatory cytokines IL-1β and IL-6 (79). CRP is elevated in AD brain (80), but is not consistently higher in AD plasma. In the current study, CRP was detected in the 2DGE but when measured in the validation cohort by LC-MS the CRP levels were sporadic and no significant elevation was observed in AD (Figure 6). The highly variable nature of the CRP levels is consistent with its role in acute phase inflammation response that changes with a large range of pathologies and not specific for AD.

Zinc α 2-glycoprotein (ZAG) is a 41 kDa glycoprotein adipokine believed to function as a lipid mobilizing factor (81). In a study with a relatively wide range of body mass indices (BMI), serum ZAG discriminated groups with a negative correlation between BMI and ZAG concentrations (82). ZAG has been found significantly lower in ventricular CSF of AD patients, discriminating AD from normal controls and non-AD dementias (83). Lower BMI has been associated with risk of AD (84). In the current study, BMI data were available for 54/72 AD subjects and 60/72 control subjects and was significantly higher in the AD pools (Table 1), consistent with elevated ZAG variants observed. The validation study on individual samples showed a trend (p=0.06) towards elevated levels of ZAG in plasma from AD cases (Figure 6).

In addition to disease classification, participants were evenly pooled according to sex. A consensus has yet to be reached as to the prevalence of sex-specific differences in AD (85,86) and there is limited proteomic data on plasma investigating sex specific differences between AD and controls. To look for sex-specific dysregulation of proteins, we made intra-sex comparisons between AD and HC pools and found Vitamin D binding protein (VDBP) and Complement factor I (CFI) to be differentially regulated according to sex. VDBP is an abundant, monomeric 52-59 kDa glycoprotein that functions as the primary plasma transporter of the sterol vitamin D and its metabolites (87). In addition to its role as vitamin D transporter, VDBP scavenges and sequesters actin monomers in the blood after cellular damage. It does so in concert with gelsolin (88), protecting micro-vasculature from the detrimental effects of ectopic actin fibrillization. VDBP is a regulator of macrophages and osteoclasts, and as an important chemotactic factor for leukocytes (89), independent of its vitamin D binding function. Its synthesis is estrogen dependent (90) and blood concentration is significantly lower in males than females (91,92). The gel image of F3 shows three abundant spot-trains of cleaved VDBP species (Figure 1 F3, arrows n, o, x), below the intact VDBP spot-train (Figure 1 F3, arrow m). The three spot-train clusters of cleaved peptides focused as disulfide bound complexes in the IEF step, to similar pIs as the uncleaved variants, as they were unified by 14 intra-chain disulfide bridges. After focusing, chemical reduction released cleaved peptides to migrate to their respective lower apparent molecular weights. The proteoform difference observed with 2D-gels was not recapitulated with the LC-MS/MS assay that measured the total pool of VDBP (Figure 6).

CFI is a complement activation inhibitor that has been shown to undergo disrupted function *in vitro* in the presence of Aβ (93). CFI plasma concentration is modestly decreased in AD and correlated with brain volume (29), but we found no reports of gender specificity. Further, when measured in individuals we did not see any gender differences.

The etiology of AD, as for many diseases, is complex and polygenic. Pooled proteomics studies are designed to detect sporadically changing disease-related isoforms that change consistently enough in afflicted subjects to effect consistent “average” changes within pools, and as such constitutes a limitation of the current study. Our future studies aim to validated and understand the relationships between VDBP, CFI, α1ACT, and α1AT with sex and genotype, respectively. Here we measured the biomarkers discovered with 2DGE using a quantitative LC-MS/MS assay and the changes we observed by 2DGE were not validated with this approach. However, we did measure samples by quantitative LC-MS/MS that were independent of the discovery study and differences between AD and control values of several of the markers approached statistical significance (e.g. VDBP; Figure 6). Overall, the measurement of total protein by LC-MS rather than proteoform-specific changes measured via 2DGE offers a potential explanation for the failure to validate the 2DGE discoveries. However, the replication of elevation of beta-2-microglobulin, complement C3 and peroxiredoxin 2 validate previous reports of these changes in AD and of the LC-MS/MS based assay to report global protein level changes (29,52-54). The correlation α1AT or α1ACT with the *APOE* genotype indicates that the point mutation differences between APOE ε2/3/4 may have wider impact than on the function of just the ApoE protein as a recent proteomic study in AD brain tissue has demonstrated (55). Further, the failure to validate many of the protoeform changes highlights the inability of assays that measure the total abundance of a protein target (e.g. ELISA, LC-MS/MS) to reflect the complexity of the proteome. Overall, complement C3, beta-2-microglobulin and peroxiredoxin-2 may add value to the emerging plasma amyloid b2 and tau assays for the early diagnosis of AD.

## Data Availability

The proteomics data used to support the findings of this study are available from the corresponding author upon reasonable request.

## Abbreviations

α1AT: α1-Anti-trypsin
α1ACT: α1 Anti-chymotrypsin
APR: Acute-phase reactant
AD: Alzheimer’s disease
APP: Amyloid precursor protein
Aβ: Amyloid β peptide
ApoE: Apolipoprotein E protein
BBB: Blood-brain barrier
BMI: Body mass index
CRP: C-reactive protein
CNS: Central Nervous System
CSF: Cerebrospinal fluid
Hpt: Haptoglobin
HC: Healthy control
HRG: Histidine-rich glycoprotein
ITIH: Inter-α trypsin inhibitor
Isoelectric point pI: Isoelectric focusing IEF
KPI: Kunitz-type protease inhibitor
RP-HPLC: Reverse phase high pressure liquid chromatography
VDBP: Vitamin D binding protein
ZAG: Zinc α 2-glycoprotein

## Acknowledgments

We would like to acknowledge the Australian Imaging and Biomarker, Lifestyle (AIBL) research team (a complete list of researchers can be found at www.aibl.csiro.au) and the volunteers and their families. We would also like to acknowledge funding from the Dementia Collaborative Research Centres (DCRC), The Victorian Government’s Operational Infrastructure Support Program and the Wicking Trust. National Institutes of Health-Small Business Technology Transfer (NIH STTR grant 5R42RR021790, EAD PI), Center for Biological Research Excellence (NIH CoBRE grants 1P20RR024237 and 2P20GM 104935, EAD PI), Montana Board of Research and Commercialization Technology Grants: (MBRCT: 05-14, 06-46, and 08-17, EAD PI) and the Murdock Charitable Trust (EAD, PI). The Cooperative Research Centre for Mental Health, Alzheimer’s Drug Discovery Foundation grant (BRR, PI), Knott Family Equipment Grant (BRR, PI), the Pierce Armstrong Trust (BRR, PI), NHMRC Dementia Leadership Fellowship (BR, APP1138673), and NHMRC project grant (BR, APP1164692). The Florey Neuroproteomics Facility. We would like to thank the Melbourne Mass Spectrometry and Proteomics Facility of The Bio21 Molecular Science and Biotechnology Institute at The University of Melbourne for the support of mass spectrometry analysis.

## Data availability statement

The proteomics data used to support the findings of this study are available from the corresponding author upon request.

